# PROMISE (Program Refinements to Optimize Model Impact and Scalability based on Evidence): A cluster-randomized, stepped-wedge trial assessing effectiveness of the revised versus original Ryan White Part A HIV Care Coordination Program for patients with barriers to treatment

**DOI:** 10.1101/19012427

**Authors:** Mary K. Irvine, Bruce Levin, McKaylee Robertson, Katherine Penrose, Jennifer Carmona, Graham Harriman, Sarah Braunstein, Denis Nash

## Abstract

**Introduction:** Growing evidence supports combining social, behavioral and biomedical strategies to strengthen the HIV care continuum. However, combination interventions can be resource-intensive and challenging to scale up. Research is needed to identify intervention components and delivery models that maximize uptake, engagement and effectiveness. In New York City (NYC), a multi-component Ryan White-funded medical case management intervention called the Care Coordination Program (CCP) was launched at 28 agencies in 2009 to address barriers to care and treatment. Effectiveness estimates based on >7,000 clients enrolled by April 2013 and their controls indicated modest CCP benefits over ‘usual care’ for short- and long-term viral suppression, with substantial room for improvement.

**Methods and analysis:** Integrating evaluation findings and CCP service-provider and community-stakeholder input on modifications, the NYC Health Department packaged a Care Coordination Redesign (CCR) in a 2017 request for proposals. Following competitive re-solicitation, 17 of the original CCP-implementing agencies secured contracts. These agencies were randomized within matched pairs to immediate or delayed CCR implementation. Data from three nine-month periods (pre-implementation, partial implementation and full implementation) will be examined to compare CCR versus CCP effects on timely viral suppression (TVS, within four months of enrollment) among individuals with unsuppressed HIV viral load newly enrolling in the CCR/CCP. Based on estimated enrollment (n=824) and the pre-implementation outcome probability (TVS=0.45), the detectable effect size with 80% power is an odds ratio of 2.90 (relative risk: 1.56).

**Ethics and dissemination:** This study was approved by the NYC Department of Health and Mental Hygiene Institutional Review Board (IRB, Protocol 18-009) and the City University of New York Integrated IRB (Protocol 018-0057) with a waiver of informed consent. Findings will be disseminated via publications, conferences, stakeholder meetings, and Advisory Board meetings with implementing agency representatives.

**Trial registration:** Registered with ClinicalTrials.gov under identifier: NCT03628287, Version 2, 25 September 2019; pre-results.

**ARTICLE SUMMARY:** *Strengths and limitations of this study:*

- The PROMISE trial, conducted in real-world service settings, leverages secondary analyses of programmatic and surveillance data to assess the effectiveness of a revised (CCR) versus original HIV care coordination program to improve viral suppression.
- To meet stakeholder expectations for rapid completion of the CCR rollout, the study applies a stepped-wedge design with a nine-month gap between implementation phases, prompting use of a short-term (four-month) outcome and a brief (five-month) lead-in time for enrollment accumulation.
- Randomization is performed at the agency level to minimize crossover between the intervention conditions, since service providers would otherwise struggle logistically and ethically with simultaneously delivering the two different intervention models to different sets of clients, especially given common challenges related to reaching agreement on clinical equipoise.^1–3^
- The use of agency matching, when followed by randomization within matched pairs, offers advantages akin to those of stratified random assignment: increasing statistical power in a situation where the number of units of randomization is small, by maximizing equivalency between the intervention and control groups on key observable variables, thus helping to isolate the effects of the intervention.^3^
- In addition, nuisance parameters are removed through the conditional analytic approach, which accounts and allows for the unavoidably imperfect matching of agencies and arbitrary variation of period effects across agency pairs.^4^

## INTRODUCTION

Successful HIV treatment at the individual level requires consistent adherence to antiretroviral therapy (ART) resulting in sustained suppression of HIV-1 viral load (VL) in plasma to levels below the detection limit of HIV RNA tests used by healthcare providers. At the population level, viral suppression (VS) is the key to the dual goals of improving health and survival among people with HIV (PWH) and preventing HIV transmission. A growing consensus among researchers supports the blending of evidence-based social, behavioral and biomedical strategies (i.e., ‘combination interventions’) to address barriers to ART use and adherence.^5–10^ However, combination interventions can be resource-intensive to deliver and challenging to translate to new and diverse organizational settings.^11^ Research is needed to systematically inform the selection of intervention components and service delivery approaches that can maximize program uptake, engagement, and effectiveness at scale in a range of practice environments.

In New York City (NYC), a multi-component Ryan White Part A–funded medical case management intervention known as the HIV Care Coordination Program (CCP) was launched in late 2009 to meet the needs of individuals with suboptimal HIV care outcomes or new HIV diagnoses. By late 2013, an NYC Health Department-City University of New York (CUNY) research partnership had secured funding for the “CHORDS” (<u>C</u>osts, <u>O</u>utcomes and <u>R</u>eal-word <u>D</u>eterminants of <u>S</u>uccess in HIV Care Coordination) study of CCP effectiveness (R01 MH101028). Early findings of increased care retention^12^ led the CDC to include the CCP in their Compendium of Evidence-based Interventions.^13^ Observational studies with a rigorously selected usual-care comparison group^14^ have shown positive CCP effects on short-term VS (Relative risk [RR] = 1.32, 95% CI:1.23-1.42),^15^ as well as durable VS (DVS) (RR: 1.16, 95% CI: 1.04-1.29),^16^ among PWH without previous evidence of VS. However, substantial room for improvement remains: over one-third of clients drop out of the program in the first year;^12,16^ and a minority of clients without previous evidence of VS achieve VS (43%)^15^ or DVS (21%).^16^ In addition, some of the original CCP design features have curbed client and provider engagement: a rigid system of program tracks has impeded service intensity adjustment based on client need (i.e., differentiated care^17,18^); a complex reimbursement model has diverted staff time and attention to maintaining agency cash flow; and a requirement for weekly visits over a three-month “induction period” for new clients has reportedly deterred eligible PWH from enrolling and discouraged staff from suggesting the CCP for PWH perceived as unable to meet that commitment.^19^

After several years of CCP implementation, the NYC Health Department and the local Ryan White Part A community Planning Council outlined a set of program modifications in response to the identified implementation barriers^19^ and the evolving epidemiologic data and intervention literature, including findings from the CHORDS research collaboration.^15,16^ Program revisions were integrated into the Health Department’s late-2017 request for proposals (RFP) initiating a competitive selection process for future NYC Care Coordination service delivery contracts. This RFP also outlined plans for agency-level randomization to an early or delayed start of the revised model, as part of an experimental evaluation of its effectiveness.

This paper describes the experimental protocol for the study known as PROMISE (<u>P</u>rogram <u>R</u>efinements to <u>O</u>ptimize <u>M</u>odel Impact and <u>S</u>calability based on <u>E</u>vidence), which continues the NYC Health Department-CUNY research partnership initiated with CHORDS. Our purpose is to inform practice-driven intervention research, particularly in the context of generating evidence for the optimization of safety-net service delivery strategies. The overarching goal of PROMISE is to investigate the impact and implementation of empirically driven course corrections to an already effective intervention model. We will test the combined effect of intervention modifications in a cluster-randomized controlled trial applying a cross-sectional, stepped-wedge design to the rollout of the revised model in previously funded, re-awarded CCP provider agencies. Drawing upon an implementation science framework and RE-AIM^20,21^ principles, we posit that the model revisions will reduce logistical and administrative barriers to service delivery and increase program engagement (among staff and clients), reach, fidelity and effectiveness. Specifically, we hypothesize that a higher proportion of PWH with unsuppressed VL at enrollment in the Care Coordination *revised* (CCR) program will achieve timely VS, as compared to PWH with unsuppressed VL at enrollment in the *original* CCP during the same period.

## METHODS AND ANALYSIS

### Participants, intervention, and outcomes

#### Study setting

Of the original 28 agencies funded to deliver the CCP since December 2009, 17 secured funding under the 2017-2018 re-solicitation, including a range of organization types (public and private hospitals, community health centers and community-based social-service organizations). Together, the 17 re-awarded agencies covered all NYC boroughs/counties and a collective caseload of more than 2,000 active CCP clients as of the end of March 2018 (agency median: 96, inter-quartile range [IQR] 82-181). Study sites are listed at: https://clinicaltrials.gov/ct2/show/NCT03628287. Agencies assigned to delayed CCR implementation received contract extensions to continue service delivery under the original CCP, while awaiting their transition to new/CCR contracts.

#### Eligibility criteria (for clients and sites)

PWH eligible for the trial analysis of CCR intervention effects include those newly enrolled in the CCP or CCR and having unsuppressed VL (HIV RNA ≥200 copies/mL) as of their latest test in the year prior to enrollment or having no VL test result reported to surveillance in the year prior to enrollment (presumed out of care^14^). To allow four months of VL outcome observation per enrollee prior to the start of the next transition (so that each outcome measure represents a single implementation phase), the trial-eligible enrollment window for each nine-month implementation period is restricted to the first five months. The trial excludes newly awarded (CCP-naïve) agencies and includes only the 17 re-awarded agencies, which could be assigned to continue CCP delivery uninterrupted or begin CCR delivery in the initial implementation phase.

#### Intervention (CCR) and control (CCP) conditions

The CCP has been described previously.^12,22,23^ The control condition is the site-level continuation of CCP model delivery, while the intervention condition is a site-level change to deliver the CCR. Sites’ ability to begin CCR delivery hinges on reimbursement for CCR services and approaches, and on receipt of Health Department trainings related to programmatic changes. Study assignments have been maintained through the scheduling of agencies for CCR trainings and contract starts (tied to the change in reimbursement).

Specific program revisions^24^ (added, changed or removed components) and their anticipated benefits are displayed in **Table 1**. Major *additions* include a training and set of tools for baseline and quarterly assessment and counseling around client HIV self-management capacity;^25^ allowance of video chat as a delivery mode for certain services, such as directly observed therapy (DOT); and optional iART^26,27^ (“immediate” ART: ensuring that the client has a filled prescription within 4 days of either enrollment or diagnosis). *Changes* include the replacement of per-member-per-day reimbursement with a fee-for-service reimbursement model that accounts for resource demands, such as staff travel to and from clients’ homes, and offers higher rates for meeting performance standards (e.g., same-day prescription fulfillment). Other *changes* include greater emphasis and guidance on identifying and recruiting individuals with documented clinical need (e.g., unsuppressed VL). Some CCP requirements (i.e., induction period, enrollment tracks) were selected for *removal* in favor of flexibility for client-centered/differentiated care.^17,18^

**Table 1.** Revisions expected to improve uptake, fidelity, engagement, effectiveness and reach/impact

|  | Added Components |  |  | Changed |  | Removed |
| --- | --- | --- | --- | --- | --- | --- |
|  | Self-manage-ment assessment | Use of video chat tools (optional) | iART (optional) | Eligibility criteria | Payment structure | Rigid program tracks |
| Uptake (provider) |  |  |  |  |  | X |
| Fidelity (provider ) |  | X |  |  | X | X |
| Engagement (client) | X | X |  |  |  | X |
| Effectiveness | X | X | X |  | X | X |
| Reach/impact | X | X | X | X | X | X |

Individual clients may switch from one of the 17 sites to another, potentially changing their intervention condition. However, based on intent to treat, clients count toward the trial only in their first enrollment during the study period. Clients in the trial may access other interventions within CCP/CCR settings or other agencies where they receive services. The NYC HIV-related services landscape includes many provider agencies and funding sources beyond those that can be tracked by any single entity; it is not feasible to prohibit simultaneous receipt of other interventions that may affect the study outcome.

#### Outcome measurement

To assess the clinical benefit of the programmatic revisions differentiating the CCR from the CCP, we will analyze client-level, surveillance-based laboratory test data.^14^ The outcome, timely VS (TVS), is defined as VL <200 copies/mL on the last VL test reported to the NYC HIV surveillance registry in the four months following CCP/CCR enrollment (TVS=1). Consistent with our prior CCP work,^12,14,15,22^ those without any VL measure during follow-up will be considered not to have achieved VS (TVS=0), given their lack of documented clinical monitoring since their last unsuppressed VL.

#### Timeline

**Figure 1** illustrates the three nine-month periods used in the stepped-wedge design: Period 0, with CCP at all 17 agencies and no CCR; Period 1, representing CCR implementation only at sites randomized to an early start (and thus encompassing the months of simultaneous operation of the CCP and the CCR); and Period 2, representing CCR implementation at all 17 sites.

**Figure 1.**
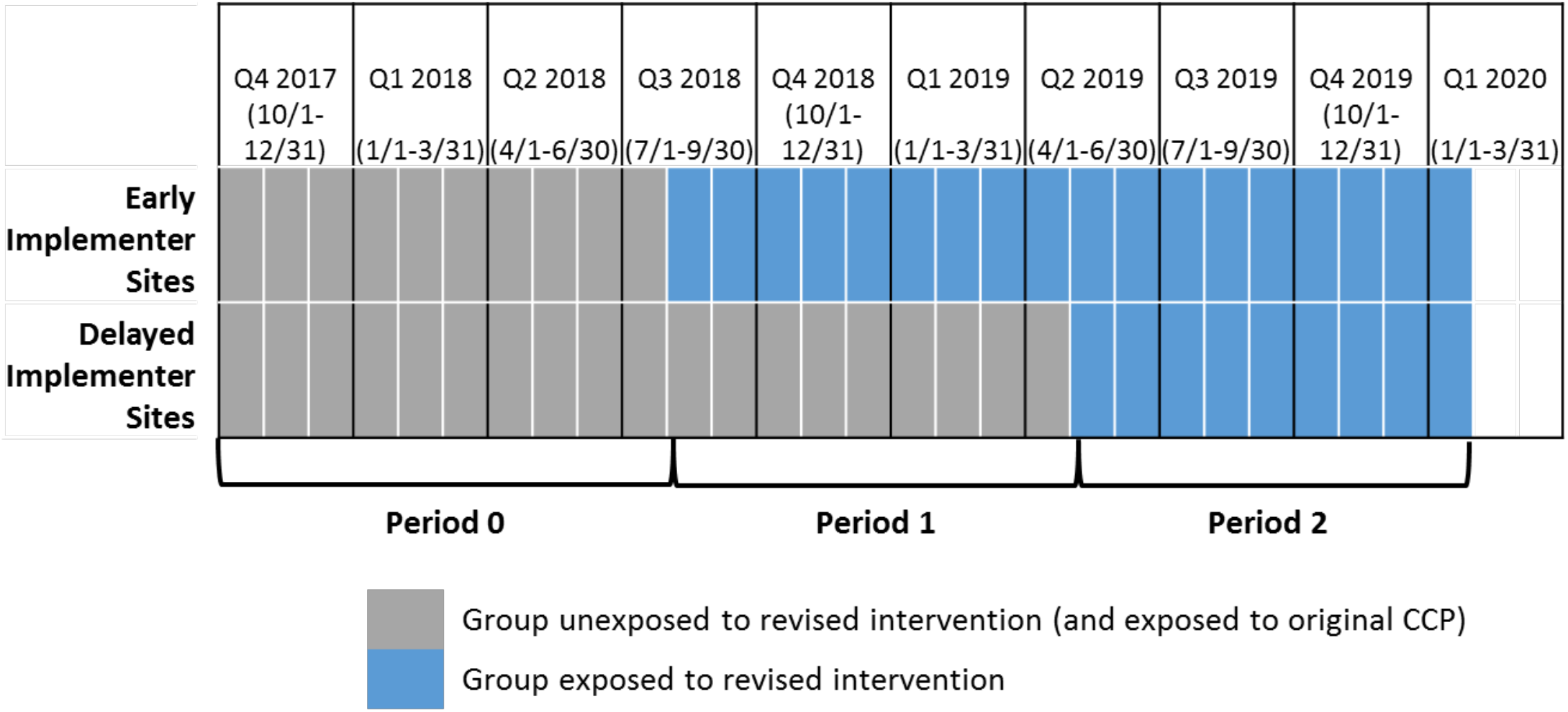
Stepped-wedge design with three implementation periods.

#### Recruitment

Beyond standard contract startup deliverables based on early program enrollment milestones, no specific incentives have been used to encourage recruitment. As informed consent has been waived for this trial, analyses will include all eligible enrollments in CCP/CCR services at any of the 17 study sites.

### Assignment of interventions

#### Randomization

Though the unit of analysis for TVS is the individual, the unit of randomization is the Care Coordination provider agency (i.e., cluster). Characteristics and study arm assignments of the 17 agencies are shown in **Table 2**. Agencies were matched and randomized within pairs (including one case in which two smaller agencies were matched to a larger one). Matching accounted for characteristics plausibly related to the TVS outcome: agency type, primary location/borough and program size (measured via a combination of CCP caseload at time of re-award and award amount). While randomization could not feasibly be stratified by each of these variables, the lead analyst suggested pairs maximizing similarity on these variables. Pairings were finalized with input from other team members knowledgeable about the programs/agencies involved. The lead analyst used a random number generator in Excel to determine agency assignments within pairs, and assignments were communicated as contract conditions in the notifications of awards.

**Table 2.** Agency characteristics, pairings and study arm assignments

| Agency ID | Award increased >20% from prior year? | Typical (prior) Caseload | Borough | Type of Site | Pair | Phase (study arm) |
| --- | --- | --- | --- | --- | --- | --- |
| 21 | Yes | 84 | Bronx | CBO/no clinical services | 1 | 1 |
| 1 | No | 101 | Bronx | CBO/no clinical services | 1 | 2 |
| 20 | Yes | 109 | Brooklyn | Public Hospital | 2 | 1 |
| 14 | No | 151 | Brooklyn | Public Hospital | 2 | 2 |
| 28 | Yes | 87 | Brooklyn | Private Hospital | 3 | 1 |
| 24 | No | 96 | Brooklyn | Community Health Center | 3 | 2 |
| 25 | No | 62 | Manhattan | Community Health Center | 4 | 1 |
| 9 | No | 78 | Manhattan | Community Health Center | 4 | 2 |
| 23 | No | 228 | Manhattan | Private Hospital | 5 | 1 |
| 18 | No | 220 | Manhattan | Private Hospital | 5 | 2 |
| 13 | Yes | 82 | Bronx | Public Hospital | 6 | 1 |
| 11 | Yes | 82 | Queens | Public Hospital | 6 | 2 |
| 5 | No | 202 | Bronx | Private Hospital | 7 | 1 |
| 4 | No | 181 | Manhattan | Private Hospital | 7 | 2 |
| 8 | Yes | 77 | Staten Island | CBO/no clinical services | 8 | 1 |
| 16 | No | 63 | Brooklyn | Community Health Center | 8 | 1 |
| 2 | No | 184 | Manhattan | Community Health Center | 8 | 2 |

#### Blinding

Blinding was not feasible for this study. Assignments were transparent to implementing agencies, study team members and interested stakeholders, since contracts are publicly available information.

### Data collection, management, and analysis

#### Data collection

As with prior studies of CCP effectiveness,^12,14–16,22^ the outcome measure for clients in both study arms will be derived from the NYC HIV surveillance registry (“the Registry”), a population-based data source of electronically reported longitudinal laboratory (VL, CD4) records on all diagnosed NYC PWH.^28,29^ Use of the Registry allows near 100% ascertainment of VS for PWH in NYC HIV medical care, regardless of specific NYC medical provider, and for periods extending before and after program enrollment or discontinuation.

Each client’s CCP/CCR enrollment agency and start date will be determined from a database of contractually required Ryan White Part A provider reporting to the Health Department, the Electronic System for HIV/AIDS Research and Evaluation (eSHARE). These program reporting-based measures are available (non-missing) for all CCP/CCR clients and all implementing agencies. Program data collection forms are located on the NYC Health Department website (https://www1.nyc.gov/site/doh/health/health-topics/aids-hiv-care-coord-tools.page).

#### Data management and quality assurance

All data for the trial are entered as part of established, legally or contractually required reporting, and are protected according to CDC physical and electronic data security and confidentiality policies.^30^ Health Department staff clean and freeze surveillance datasets on a quarterly basis, and conduct matches of program to surveillance data semi-annually, with human review of each near-match by two independent analysts and a separate “tie-breaker” when the analysts’ determinations differ. Details on the deterministic matching algorithm have been previously described.^31^ Through the match, participants are assigned a unique record number used in merging surveillance and programmatic data for analytic datasets, which are stripped of all personal identifiers prior to analysis and stored on the most secured drives on the Health Department network. eSHARE data quality is checked by Health Department analysts at the time of each monthly extraction. For purposes of payment, provider agencies also review draft extracts and fill in any missing enrollment and services data monthly.

#### Statistical analysis for the matched-pairs stepped-wedge trial

##### Analysis overview and rationale

We will apply an innovative, fully conditional analysis that, in addition to allowing for arbitrary period effects, allows for arbitrary within-pair site differences. The analysis plan is based on the exact, conditional distribution theory of non-central multiple hypergeometric distributions and their convolutions,^4^ which will enable us to estimate and test the effect of the revised intervention as a single parameter defined below. The conventional statistical analysis proposed for cross-sectional stepped-wedge designs (i.e., with independent samples of clients enrolled at each transition step)^32^ assumes a mixed model with random cluster effects and fixed period effects, but this is not appropriate for our matched-pairs stepped-wedge trial. For one, the matching of pairs is under the investigators’ control and so should be conditioned upon. Second, the generalized linear mixed model has limitations, such as a gratuitous and unverifiable assumption of normal distribution for random effects and poor variance estimation performance in small samples (of clusters), even with robust variance estimation, such that jackknifing must be used. However, the following exact analysis avoids those problems by conditioning out the nuisance parameters.

##### Analysis approach and assumptions

As shown in **Table 3**, for each pair of sites, we will produce two 2×3 tables (one table per site in pair), cross-classifying the number of TVS and non-TVS outcomes in Period 0 (with original CCP but no CCR implementation), Period 1 (first transition, with CCR only at sites assigned to an early start), and Period 2 (second transition, with CCR at all sites). For identification purposes, we refer to “Site 1” within a matched pair as the site randomized to switch in Period 1 (early start) and “Site 2” as the site randomized to switch in Period 2 (delayed start). We begin by assuming the following logistic regression model for the three binomial outcomes: the logit of the probability of TVS for a given site, period and intervention (CCR versus original CCP) equals an intercept representing an arbitrary, pair-specific log odds on TVS for Site 2 in the pair, plus an arbitrary log odds ratio (LOR) for Site 1 versus Site 2 in the pair (allowing for imperfectly matched sites), plus two arbitrary pair-specific LORs for Period 1 and Period 2 effects relative to Period 0, plus one structural LOR of interest, the global intervention effect (non-existent in Period 0, applicable to Site 1 in Period 1, and applicable to both sites in Period 2). The exponent of this last parameter is the target of statistical inference, namely, the odds ratio (OR) for TVS versus non-TVS comparing the CCR to the CCP. A key assumption is that any site effects apply in each period and any period effects apply to each site, independent of the intervention effect (i.e., that there are no site-by-intervention or period-by-intervention interactions). This assumption will be tested and the model elaborated if needed. Note that under the key assumption, the constant site and period effects are allowed to vary arbitrarily from one matched pair to the next.

**Table 3.** Illustration of 2×3 tables cross-classifying TVS and non-TVS outcomes by period, for the estimation of the CCR intervention effect.

|  |  | Period 0 | Period 1 | Period 2 | Total |
| --- | --- | --- | --- | --- | --- |
| Site 1 in pair *i*<br>(adopts CCR<br>in Period 1) | TVS | $X_{i10}$ | $X_{i11}$ | $X_{i12}$ | $X_{i1+}$ |
| | No TVS | $N_{i10} - X_{i10}$ | $N_{i11} - X_{i11}$ | $N_{i12} - X_{i12}$ | $N_{i1+} - X_{i1+}$ |
| | Total | $N_{i10}$ | $N_{i11}$ | $N_{i12}$ | $N_{i1+}$ |
| Site 2 in pair *i*<br>(adopts CCR in<br>Period 2) | TVS | $X_{i20}$ | $X_{i21}$ | $X_{i22}$ | $X_{i2+}$ |
| | No TVS | $N_{i20} - X_{i20}$ | $N_{i21} - X_{i21}$ | $N_{i22} - X_{i22}$ | $N_{i2+} - X_{i2+}$ |
| | Total | $N_{i20}$ | $N_{i21}$ | $N_{i22}$ | $N_{i2+}$ |
| Pair *i* totals | TVS | $X_{i+0}$ | $X_{i+1}$ | $X_{i+2}$ | $X_{i++}$ |
| | No TVS | $N_{i+0} - X_{i+0}$ | $N_{i+1} - X_{i+1}$ | $N_{i+2} - X_{i+2}$ | $N_{i++} - X_{i++}$ |
| | Total | $N_{i+0}$ | $N_{i+1}$ | $N_{i+2}$ | $N_{i++}$ |

Note: The red font indicates the two 2x3 tables in site pair *i*. The blue font indicates the margins upon which the analysis will condition. Margins fixed by summing or subtracting other fixed margins (e.g., $N_{i1+}$ and $X_{i++}$ ) are indicated in purple font.

##### Estimating the CCR intervention effect

Next, by conditioning on the marginal totals within each site (numbers of eligible clients enrolled in each period and total numbers of TVS and non-TVS outcomes for each site), the joint distribution of the numbers of TVS outcomes for Site 1 by period becomes a non-central multiple hypergeometric distribution with only three parameters (the period LORs and the intervention LOR); i.e., the conditional distribution does not depend on the nuisance site parameters. By further conditioning on the sum of TVS outcomes across the two sites in each period, the fully conditional joint distribution depends on only one parameter, the intervention effect; i.e., the fully conditional distribution depends neither on the nuisance site effects nor on the nuisance period effects. In fact, the sufficient statistic for the intervention LOR in the fully conditional likelihood function is simply the number of TVS outcomes from Site 1 in Period 1. It is then straightforward to calculate the marginal distribution of this outcome as a function of the intervention effect. Therefore, we will calculate that distribution for each of the 8 matched pairs (including the case of two programs jointly matched to a third) and convolute those distributions to obtain the sampling distribution of the sum of sufficient statistics. Once we obtain the fully conditional sampling distribution of the sufficient statistic as described above, we will report the conditional maximum likelihood estimate of the intervention LOR with an exact, test-based 95% confidence interval. The test of the null hypothesis at the two-tailed 0.05 significance level will be based on the exact two-tailed *P*-value (using the point probability definition),^4^ and will form the primary outcome analysis. In sensitivity analyses, we will also report the Wald, Score and Likelihood Ratio test results, which should be close to each other, given expected client numbers per site per period and the level of TVS from baseline CCP data.

### Sample size

For the planning of the study, we used April 2012 – June 2014 CCP data to provide a set of marginal totals and proportions of TVS in 9 matched pairs of sites. We then prepared a simulation study with 10,000 replications to estimate the power of the primary test of intervention effect. For any given simulation replication, each site within the nine matched pairs was randomly assigned to transition at Period 1 or Period 2 (with independent randomizations per replication). For the site and period effects, we used the actual past Period 0 CCP data for the two sites to provide the within-pair site effect and the (randomly selected) second site’s TVS proportions in Period 1 and Period 2 to provide the period effects. We then applied a given intervention effect to Site 1 in Period 1 and to both sites in Period 2, for a set of plausible TVS proportions. For each such replication, we recorded the results of the exact conditional analysis described above. The pre-randomization power of the primary test was estimated as the proportion of exact two-tailed *P*-values ≤0.05.

The resulting detectable effect size (80% power with exact Type I error rate ≤0.05 two-tailed) was an OR of ∼2.15. Since odds ratios overstate risk ratios (RRs) when the outcome proportion is common, to aid in interpretation, **Table 4** indicates what detectable revised-CCP TVS proportions and RRs would correspond to various base proportions. The final two columns indicate what the power would have been for various other intervention-effect ORs. In summary, the planned study had a detectable OR of 2.15, corresponding to RRs between 1.37 and 1.53. Power estimates ranged between ∼73% and 85% for true ORs between 2.00 and 2.25, respectively.

**Table 4.** Power calculations for the Care Coordination Redesign effect on TVS (as originally planned)

| Reference<br>P [TVS] | Detectable<br>P [TVS] | Risk ratio at<br>Detectable P [TVS]<br>for True OR=2.15 | True<br>OR | Power<br>(%) |
| --- | --- | --- | --- | --- |
| 0.35 | 0.537 | 1.53 | 2.25 | 84.8 |
| 0.40 | 0.589 | 1.47 | 2.20 | 83.4 |
| 0.45 | 0.638 | 1.42 | 2.15 | 80.4 |
| 0.50 | 0.683 | 1.37 | 2.10 | 78.1 |
| nb: Average P[TVS] among all sites in all<br>periods = 0.437. Monte Carlo standard error<br>for power values is less than 0.5%. |  |  | 2.05 | 75.6 |
|  |  |  | 2.00 | 72.8 |

**Table 5** provides the corresponding detectable effect size and power values given current actual, post-randomization numbers of eligible enrollees for Periods 0 (N=169) and 1 (N=389), a conservative estimate of eligible enrollees for Period 2 (N=266), and assumed TVS proportions based on actual proportions for Period 0. The table reflects lower enrollments than planned and the randomization of eight rather than nine matched pairs. Because the randomization of sites within pairs has already been set, the simulations for Table 5 condition on this fact; i.e., early- and late-implementing sites are considered fixed as randomized. The post-randomization power is somewhat greater than the pre-randomization power, and this partially offsets the decrease in power due to lower-than-expected enrollments. The detectable effect size (80% power with exact Type I error rate ≤0.05 two-tailed) is currently an OR of 2.90, corresponding to RRs between 1.49 and 1.74. Power estimates now range between ∼76% and 83% for true ORs between 2.75 and 3.00, respectively.

**Table 5.** Power calculations for the Care Coordination Redesign effect on TVS (as currently estimated)

| Reference P [TVS] | Detectable P [TVS] | Risk ratio at Detectable P [TVS] for True OR=2.90 | True OR | Power (%) |
| --- | --- | --- | --- | --- |
| 0.35 | 0.610 | 1.74 | 3.00 | 82.8 |
| 0.40 | 0.659 | 1.65 | 2.95 | 81.1 |
| 0.45 | 0.704 | 1.56 | 2.90 | 80.6 |
| 0.50 | 0.744 | 1.49 | 2.85 | 78.7 |
| nb: Average P[TVS] among all sites in base period = 0.465. Monte Carlo standard error for power values is less than 0.5%. |  |  | 2.80 | 77.8 |
|  |  |  | 2.75 | 75.8 |

### Patient and public involvement

Starting in 2016, the local HIV Planning Council (comprising practitioners, patients and researchers), as well as Care Coordination service providers not on the Council, co-developed the revised intervention model with Health Department representatives. The plan for this trial was communicated to the Planning Council co-chairs, who provided letters of support in September 2017 after receiving the proposal aims. During a September 2017 public ‘town hall’ meeting to discuss the redesign and re-solicitation, Health Department staff described the intent to use phased implementation with random assignments for a side-by-side comparison of the two models. The approach was further outlined in the December 2017 RFP, which incorporated community feedback. The community was not involved in the methods for agency matching and randomization, due to timing and potential conflicts of interest. Once awarded, the study team engaged six implementing service agencies as study partners and began Advisory Board meetings with those partners, who will advise on dissemination of findings and have contributed to instrument design and recruitment planning for the relevant parts of the larger study. There is no recruitment or primary data collection specifically for the trial.

### Monitoring

The Health Department will oversee data monitoring and protocol compliance. No more than minimal risk is attached to study participation, which involves receipt of one model or another similar model of support services, rather than any medical device or treatment. As the intervention is delivered by contracted agency staff, and the analysis is based on secondary data sources, study investigators have no direct contact with human subjects for the purpose of the trial. To enroll in NYC HIV Care Coordination services, participants sign a consent form that covers the uses of data applicable to this trial. Given the determination of minimal risk and the routine nature of secondary analyses of merged programmatic-surveillance data as part of Health Department evaluation activities, the study team will not convene a Data Safety and Monitoring Board or conduct audits.

## ETHICS AND DISSEMINATION

This trial was approved by the NYC Department of Health and Mental Hygiene Institutional Review Board (IRB) under protocol number: 18-009, and the CUNY Integrated IRB under protocol number: 018-0057, and is registered with ClinicalTrials.gov (Identifier NCT03628287). The trial was granted a waiver of informed consent in accordance with 45 CFR 46.116(d), based on its reliance on secondary data analysis. Any changes to the trial eligibility criteria, outcome measures or analysis plans would be mutually agreed upon between the CUNY and Health Department Principal Investigators, vetted with the PROMISE Advisory Board, and submitted to the IRB as protocol modifications. We anticipate no changes that would affect CCR enrollment at this stage.

The full IRB protocol and statistical code will be made available by the investigators upon request. Due to legal restrictions (New York Public Health Law Article 21, Title III) and the confidential nature of HIV surveillance data, the study team cannot release a de-identified individual-level public-use dataset. NYC Health Department staff retain sole custody of the merged study datasets and are available to assist external researchers with any inquiries. Requests can be sent via email to.

Results will be reported in accordance with the Consolidated Standards of Reporting Trials extension to cluster-randomized trials.^33^ We will disseminate results of this study through scientific conference presentations, peer-reviewed publications, and meetings with key stakeholders. Locally, results will be communicated annually at Health Department-convened meetings with CCR provider agencies, semi-annually in PROMISE Advisory Board meetings, and approximately semi-annually at public HIV Planning Council meetings. The investigators have also been sharing this work with NIH leadership,^34^ the Research Synthesis and Translation Team of the CDC Prevention Research Branch,^35^ and interventionists and researchers in other jurisdictions (e.g., Seattle-King County).

## Conclusion

Phasing in intervention implementation within a one-year period and using random agency assignment (within pairs) to early or delayed implementation offers a means of rigorously evaluating a set of changes to a major public-services program, while ensuring fair and uninterrupted access to program benefits in the eligible population. Intentionally staggered starts can also offer advantages in terms of managing the practical demands (e.g., trainings, technical assistance and administrative work) of a large-scale program rollout. Through robust health department-university partnerships that include joint planning of research in advance of key policy or practice initiatives, locally important research questions can be answered without substantially slowing the pace of desired change, and with methods that support knowledge generation and generalizability. In this case, NYC’s experience with implementing course corrections to a complex evidence-based HIV care intervention will yield findings that can valuably inform multiple jurisdictions’ efforts to advance progress along the HIV care continuum and ultimately end the epidemic.

## Author contributions

MI is the health department (DOHMH) Principal Investigator and led the conception of the overall study and the writing of the R01 proposal. BL (CU) led the conception of the matched cross-sectional stepped-wedge design and wrote the detailed statistical analysis plan. BL, MR (CUNY), KP (DOHMH) and DN (CUNY) contributed substantively to the study design and analysis plan, conducted analyses, and provided tables/figures. JC (DOHMH) contributed centrally to the Care Coordination Redesign, wrote the RFP for the re-solicitation of service delivery contracts – which functioned as a blueprint for the CCR model, oversaw technical assistance and training to CCR providers, and facilitated communications with several provider agencies partnering on the study. GH (DOHMH) contributed to the Care Coordination Redesign, oversaw Health Department communications with the local HIV Planning Council regarding this trial, and engaged health department leadership in approving the integration of a cluster-randomized, stepped-wedge trial design into the structure for the Care Coordination re-solicitation and timing of contract starts. SB (DOHMH) oversaw the preparation of all HIV surveillance-based datasets and developed relevant surveillance data use agreements. DN is the university-based Principal Investigator and shares responsibility for the conceptualization and oversight of the overall study and this trial, serves as the primary contact with the grant funding agency, and guides dissemination. MI drafted the manuscript, and all authors (MI, BL, MR, KP, JC, GH, SB, and DN) revised it for critically important intellectual content and provided final approval of the manuscript.

## Acknowledgements

The authors are indebted to Ryan White Part A HIV Planning Council members and to Care Coordination service providers, quality management specialists, and clients, for their invaluable and ongoing contributions to real-world program implementation, evolution and evaluation. We also wish to thank Kent Sepkowitz for his keen eye and helpful critiques of drafts, and Levi Waldron, Beau Mitts, Julie Myers, and Bisrat Abraham for their involvement in the earlier effectiveness R01 study, CHORDS (Costs, Health Outcomes, and Real-world Determinants of Success in HIV Care Coordination). Lastly, we wish to recognize Sarah Kulkarni, whose skills, instincts, intellect and grant management contributions have been essential to both the CHORDS and PROMISE studies of Care Coordination.

